# Optimising the use of molecular tools for the diagnosis of yaws

**DOI:** 10.1101/19000232

**Authors:** Morgan Munson, Benjamin Creswell, Kofi Kondobala, Bawa Ganiwu, Rita Dede Lomotey, Paul Oppong, Frederick Opoku Agyeman, Nana Kotye, Mukaila Diwura, Ebenezer Padi Ako, Shirley Victoria Simpson, Kennedy Kwasi Addo, Harry Pickering, Rebecca Handley, Joanna Houghton, Cynthia Kwakye, Michael Marks

**Affiliations:** Clinical Research Department, Faculty of Infectious and Tropical Diseases, London School of Hygiene & Tropical Medicine, Keppel Street, London, United Kingdom; Ghana Health Service, Accra, Ghana; Department of Bacteriology, Noguchi Memorial Institute for Medical Research, University of Ghana, Legon, Ghana; Hospital for Tropical Diseases, Mortimer Market Centre, London, United Kingdom

## Abstract

**Introduction:** Yaws is a neglected tropical disease and results in lesions of skin, soft tissues and bones. PCR plays an important part in surveillance.

**Methods:** Children suspected to have yaws were enrolled. From the largest lesion paired swabs were collected; one into transport medium and one as a dry swab. In children with multiple lesions we collected additional swabs from up to four subsequent lesions. Swabs in transport medium were maintained in a cold-chain whilst dry swabs were stored at ambient temperature. Swabs were tested by PCR for *Treponema pallidum* and *Haemophilus ducreyi*.

**Results:** Of 55 individuals, 10 (18%) had at least 1 positive PCR for *T. pallidum* and 12 (22%) had at least 1 positive result for *H. ducreyi*. Concordance was 100% between swabs in transport medium and dry swabs. One patient had PCR-confirmed yaws on the swab of a third lesion when both the first and second lesions were PCR-negative.

**Discussion:** Storing swabs in transport medium and transporting in a cold chain did not improve yield, however detection of *T. pallidum* is increased by swabbing additional lesions. As the target for yaws is eradication approaches to sample collection need revisiting to ensure cases are not missed.

## Introduction

Yaws, caused by the *Treponema pallidum* subsp. p*ertenue*, is a neglected tropical disease (NTD) and results in lesions of the skin, soft tissues and bones[1,2]. The disease predominantly affects children and is known to be endemic in 15 countries[3]. Transmission of yaws occurs from person to person through direct skin contact with infectious exudates of early skin lesions of infected individuals[4]. Early, active yaws is characterised by papillomatous and ulcerative lesions which are highly infectious. Untreated lesions of early yaws may spontaneously heal but periods of relapse to infectious yaws may continue for a number of years. In a proportion of untreated individuals the disease progresses to highly destructive lesions of the skin and bones which result in chronic disfigurement and disability[2,5].

Interest in yaws has increased in recent years, following studies showing azithromycin to be highly effective in the treatment of yaws and that community mass treatment by azithromycin reduces the prevalence of both active and latent disease[6–10], culminating in renewed efforts by the World Health Organization (WHO) to achieve global yaws eradication[11]. Despite global eradication efforts led by WHO and UNICEF between 1952 and 1965 and further national efforts between 1981 and 1983, Ghana remains endemic for yaws, and reports the most cases of any country in Africa[3,4]. Yaws remains an important public health problem in the country and is reported in all the 10 regions of Ghana and nearly all the districts in the country. In 2008 Ghana launched the National Yaws Eradication Program (NYEP). The programme has succeeded in increasing the number of contacts treated per yaws case from 3.5 in 2008 to 10.8 in 2013[12]and in conducting pilot implementations of the WHO ‘Morges’ strategy for yaws eradication[10,11]. Currently in Ghana, as in other endemic countries, the diagnosis of yaws is most often based on clinical grounds alone without the aid of serological or molecular investigations. Studies in many settings have indicated that clinical diagnosis alone may be unreliable[13–15] and WHO has strongly advocated the introduction of diagnostic tests to improve the quality of epidemiological data on yaws in endemic countries[16].

Serological methods remain the mainstay for laboratory confirmation of yaws and include both treponemal and non-treponemal tests. Treponemal tests such as the *Treponema pallidum* particle agglutination (TPPA) assay are highly specific, but remain positive for life following infection. Diagnosis of current disease therefore relies on both a positive treponemal test but also a reactive non-treponemal tests such as the rapid plasma regain (RPR) or the Venereal Disease Research Laboratory (VRDL) test. Titres of non-treponemal tests rise during infection and fall following successful treatment [2]. Rapid diagnostic tests that combine treponemal and non-treponemal components, such as the dual path platform (DPP) ‘syphilis screen and confirm’ assay, have now been validated for the diagnosis of yaws and have largely replaced traditional laboratory assays within the programmatic setting [17–20].

Polymerase chain reaction (PCR) methods to detect treponemal DNA from genital ulcers of venereal syphilis have been available for more than two decades [21,22]. More PCR assays have been developed that are able to differentiate between the DNA of different subspecies of *T*.*pallidum* complex[14,23]. Due to primer binding-site mutations occurring in some strains of yaws the current recommendation is to use a pan-treponemal PCR target such as polA or tp47 for the diagnosis of yaws[24]. When PCR is used to assess individuals with clinical and serological evidence of yaws a large proportion of lesions are found to contain no treponemal DNA[23,25]. In several studies *Haemophilus ducreyi* has been demonstrated to be a common cause of skin lesions which may be confused with those of yaws in endemic countries in the South Pacific islands and Africa [15,25,26]. For this reason there is recognition that PCR will play an increasingly important part in yaws surveillance efforts worldwide [16].

Currently there is limited data on the optimal approach to collecting samples for molecular confirmation of yaws. In a number of studies samples have been collected from a single lesion per individual, placed in transport medium and shipped within a strict cold chain[23,25,27]. A number of studies assessing the use of PCR for the diagnosis of yaws have suggested that transport medium is unnecessary and that dry swabs perform equally well[28]. A move to dry swabs and shipping at ambient temperatures would reduce programmatic costs and potentially increase feasibility of incorporating molecular testing into the programmatic context. In this study we aimed to evaluate whether collection of additional swabs, from more than a single lesion, and the presence or absence of transport medium affected the performance of PCR for the diagnosis of yaws.

## Methods

### Case Finding and Screening

The study was conducted in the West Akim and Upper West Akim districts in the Eastern Region of Ghana. These districts were selected as they are known to be highly endemic for yaws. We conducted active case finding searches in July 2018 in collaboration with the National Yaws Eradication Project (NYEP). Field workers visited schools to identify children with lesions consistent with yaws. Children with sores, ulcers or skin lesions were asked to come forward for more detailed examination as per routine NYEP protocol during case searching. Children with yaws-like ulcers were first screened with the SD Bioline syphilis test and those who tested positive were then tested with the DPP Syphilis Screen and Confirm test. Individuals with a dual positive (treponemal and non-treponemal) DPP assay were enrolled into the study.

### Swab Collection

We identified a maximum of 5 yaws-like lesions per individual prior to treatment. Swabs were taken in line with the WHO recommendations for swabbing yaws lesions. In brief, a single Dacron swab was used to obtain tissue from each lesion. This was performed by pressing the head of the swab into the centre of the lesion and rotating the swab over the area. If a lesion or papilloma was dry, the area was moistened using sterile saline on some gauze and the outermost epithelial lining was gently removed using a blunt object before swabbing. We identified the largest, most exudative lesions first. From this lesion paired swabs were collected; the first into transport medium and the second as a dry swab. From a maximum of four subsequent lesions we collected additional dry swabs at baseline. All individuals were then offered a single-dose of azithromycin in line with national guidelines. Follow-up dry swabs were collected in the same manner from the first three lesions at 8, 24, 36 and 48 hours after treatment with azithromycin.

### Swab Storage

Swabs in transport medium were placed immediately into a cool box and were frozen at −20°C at the laboratories of Noguchi Memorial Institute for Medical Research (NMIMR). Swabs in transport medium were then shipped on dry-ice to the UK where they were again frozen at - 20°C until testing. Dry swabs not in transport medium were stored at ambient temperature in the field for the duration of the study and shipped at ambient temperature to the UK for testing.

### Swab Testing

DNA was extracted from lesion swabs using the QIAamp DNA Mini Kit according to the manufacturer’s instructions. Multiplex PCR targeting the tp47 of *T. pallidum* and the hhdA gene of *H. ducreyi* was performed on a Rotorgene 3000 as previously described[22]. If baseline swabs were PCR positive for either target, the follow-up swabs from the same lesion were extracted and tested using the same PCR targets. Positive and no-template controls were included in every PCR run.

### Analysis

We compared the proportion of samples positive by PCR for either *T. pallidum* or *H. ducreyi* for the paired lesion samples collected from the ‘target’ lesion. We considered the samples collected in transport medium and maintained in cold chain as the gold standard and the dry swabs stored at ambient temperature to be the comparator. We calculated the proportion of individuals diagnosed with either *T. pallidum* or *H. ducreyi* based only on a swab of the target lesion to the proportion diagnosed based on swabbing of multiple lesions. For individuals whose lesion was PCR positive at baseline we assessed the number of lesions in which pathogen DNA remained detectable after treatment with azithromycin.

### Ethics Approval

Ethics approval for the study was obtained from the Ghana Health Services Ethics Review Committee and London School of Hygiene and Tropical Medicine. Parents or guardians provided written informed consent for children to participate in the study and where appropriate assent was also obtained from children.

## Results

Approximately 3,000 children were screened during active case searches to identify 55 individuals with clinical evidence of yaws and a dual positive DPP assay. The median age of individuals enrolled in the study was 10 years (IQR 9-12) and a majority of patients were male (n = 33, 60%). For dry swabs the median duration of storage at ambient temperature was 22 days (IQR 21-32).

Overall, 10 (18%) patients had at least 1 positive PCR result for *T. pallidum* and 12 (22%) patients had at least 1 positive PCR result for *H. ducreyi*, representing 22% of all participants. Of the 55 children included in this study, 53 had a swab from their target lesion collected both into transport medium and a dry swab at baseline. There was 100% concordance between the results of PCR conducted on swabs collected in transport medium and maintained in a cold chain and dry swabs stored at ambient temperature.

Overall 111 baseline swabs were collected from the 55 patients enrolled in the study. The median number of swabs per patient was 2 (IQR 1-2.5). Overall a total of 14 swabs (12.6%) were positive for *T. pallidum* of which 9 swabs were from target lesions. Four swabs were positive from additional lesions of patients whose target lesion was also PCR-positive for *T. pallidum*. One patient had PCR confirmed yaws diagnosed based on the swab of a third lesion when both the target and secondary swabbed lesion were PCR-negative. Overall a total of 17 swabs (15.3%) were positive for *H. ducreyi* of which 9 were from target lesions. Five swabs were positive from additional lesions of patients whose target lesion was also PCR-positive for *H. ducreyi* whilst three samples were positive from individuals whose target lesion was PCR-negative for *H. ducreyi*. Of 14 swabs that were positive at baseline for *T. pallidum*, follow-up swabs were available for 11 lesions. Of these 11 lesions every available follow-up swab remained PCR-positive, including the seven patients who had swabs collected 48 hours after treatment.

## Discussion

PCR has become an important tool in diagnosis and surveillance for yaws as part of global eradication efforts. In this study we demonstrate that storing swabs in transport medium and transporting them within a cold chain does not improve the diagnostic yield compared to dry swabs stored at ambient temperature. Our findings may have significant cost and logistical implications for programmes and should increase the feasibility of incorporating molecular diagnostics into routine care and surveillance activities. In addition, despite our relatively small sample size, we also demonstrated that the detection of *T. pallidum* could be increased by swabbing additional lesions beyond the ‘target’ lesion. Swabbing only a single lesion has been practice in all studies to date[15,23,25,27] but our data suggests such an approach will result in misclassification of cases. As the target for yaws is global eradication, detection of every case is vital and our study demonstrates that the approach to sample collection will need to be revisited to ensure cases of active yaws are not missed.

Access to PCR to confirm the diagnosis of yaws remains extremely limited in all endemic countries outside of research studies. Diagnostic confusion with *H. ducreyi* and the need to monitor for the emergence of azithromycin resistance in *T. pallidum* highlight that molecular tools will be required to support global yaws eradication efforts[16,29]. This is the first study to assess the impact of the use of transport medium and a cold-chain on the diagnostic yield of lesion swabs for PCR. The cycle threshold (CT) value of the PCR assay was also similar for both dry swabs and swabs collected in transport medium (data not shown) suggesting that DNA was also not adversely affected by the absence of transport medium or a cold-chain. Taken together with existing studies on syphilis our data suggests that there is no benefit in using transport medium or a cold chain for routine diagnostic purposes. Simplifying sample collection, storage and transport requirements should reduce cost and improve the feasibility of implementing PCR as part of programmatic activities.

Many patients with yaws have multiple lesions on examination however to date studies have focused on collecting a sample from only the largest ‘target’ lesion. Such a strategy presumes that all skin lesions in an individual are caused by the same pathogen. In this study we clearly demonstrate that this may result in missing PCR positive cases of yaws. As the aim of the WHO strategy is global eradication detection of every case is vital and our data strongly argues that consideration be given to swabbing multiple skin lesions where these are present to ensure all cases of active yaws are detected. We used separate swabs for each lesion and tested these separately. Pooling of samples for testing or use of a single swab for multiple lesions may facilitate similar increases in diagnostic yield whilst avoiding the additional cost of performing separate PCR on multiple lesions. We also demonstrated that lesions remain positive by PCR up to 48 hours after treatment. Whilst a positive PCR after treatment might simply represent detection of DNA and not viable organisms, our findings support the WHO recommendation to provide wound dressings as well as treatment to reduce any potential risk of ongoing transmission post-treatment.

The major limitation of our study is the relatively small sample size. It is possible that with a larger number of *T. pallidum* PCR positive lesions that we might have been able to demonstrate a difference in the performance of PCR between samples collected into transport medium compared to dry swabs. However, studies in syphilis have also demonstrated no difference between samples collected into transport medium or dry swabs [28,30,31]. Samples were stored at ambient temperature for a median of more than twenty days which is likely to be longer that the average duration of sample transport under programmatic conditions. We believe therefore, that our results are robust and that in routine settings there is no requirement for transport medium or a cold chain when collecting diagnostic samples. Secondly, swabbing of additional lesions detected only a single extra case of PCR confirmed active yaws. However, this additional case represented 10% of all cases of active yaws diagnosed in the study. For the global eradication campaign, it will be vital to detect every case, and our data strongly supports the need to consider swabbing multiple lesions when they are present.

## Conclusion

Molecular tests will play a central role in surveillance and monitoring during yaws eradication efforts but there have been few studies evaluating how best to operationalise this tool. By simplifying sample collection, storage and transport and highlighting the importance of sampling multiple lesions, the data from our study should enable programmes to optimise their use of this critical diagnostic tool and support successful global eradication efforts.

## Data Availability

Data is available in the supplementary file

## Statements on the authors’ contributions

MM, BC, JH, CK, MM designed the study. MM, BC, HP, RH, JH, CK, MM analysed and interpreted the data. MM, BC, CK and MM prepared the first draft of the manuscript. All authors contributed to study implementation and revision of the manuscript.

## References

1. Marks M, Solomon AW, Mabey DC. Endemic treponemal diseases. Trans R Soc Trop Med Hyg. 2014;108: 601–607. doi:10.1093/trstmh/tru128

2. Mitjà O, Asiedu K, Mabey D. Yaws. Lancet. 2013;381: 763–773. doi:10.1016/S0140-6736(12)62130-8

3. Mitjà O, Marks M, Konan DJP, Ayelo G, Gonzalez-Beiras C, Boua B, et al. Global epidemiology of yaws: a systematic review. Lancet Glob Health. 2015;3: e324–331. doi:10.1016/S2214-109X(15)00011-X

4. Asiedu K, Fitzpatrick C, Jannin J. Eradication of Yaws: Historical Efforts and Achieving WHO’s 2020 Target. PLoS Negl Trop Dis. 2014;8: e3016. doi:10.1371/journal.pntd.0003016

5. Perine PL, Hopkins DR, Niemel PLA, St. John R, Causse G, Antal GM. Handbook of endemic treponematoses⍰: yaws, endemic syphilis and pinta [Internet]. Geneva, Switzerland: World Health Organization; 1984. Available: http://apps.who.int/iris/handle/10665/37178?locale=en

6. Kwakye-Maclean C, Agana N, Gyapong J, Nortey P, Adu-Sarkodie Y, Aryee E, et al. A Single Dose Oral Azithromycin versus Intramuscular Benzathine Penicillin for the Treatment of Yaws-A Randomized Non Inferiority Trial in Ghana. PLoS Negl Trop Dis. 2017;11: e0005154. doi:10.1371/journal.pntd.0005154

7. Mitjà O, Hays R, Ipai A, Penias M, Paru R, Fagaho D, et al. Single-dose azithromycin versus benzathine benzylpenicillin for treatment of yaws in children in Papua New Guinea: an open-label, non-inferiority, randomised trial. Lancet. 2012;379: 342–347. doi:10.1016/S0140-6736(11)61624-3

8. Mitjà O, Houinei W, Moses P, Kapa A, Paru R, Hays R, et al. Mass Treatment with Single-Dose Azithromycin for Yaws. N Engl J Med. 2015;372: 703–710. doi:10.1056/NEJMoa1408586

9. Marks M, Vahi V, Sokana O, Chi K-H, Puiahi E, Kilua G, et al. Impact of Community Mass Treatment with Azithromycin for Trachoma Elimination on the Prevalence of Yaws. PLoS Negl Trop Dis. 2015;9: e0003988. doi:10.1371/journal.pntd.0003988

10. Abdulai AA, Agana-Nsiire P, Biney F, Kwakye-Maclean C, Kyei-Faried S, Amponsa-Achiano K, et al. Community-based mass treatment with azithromycin for the elimination of yaws in Ghana—Results of a pilot study. PLoS Negl Trop Dis. 2018;12: e0006303. doi:10.1371/journal.pntd.0006303

11. The World Health Organisation. Eradication of yaws - the Morges Strategy. Wkly Epidemiol Rec. 2012;87: 189–194.

12. Agana-Nsiire P, Kaitoo E, Agongo EEA, Bonsu G, Kyei-Faried S, Amponsa-Achiano K, et al. Yaws Prevalence, Lessons from the Field and the Way Forward towards Yaws Eradication in Ghana. Int Sch Res Not. 2014;2014: 1–7. doi:10.1155/2014/910937

13. Fegan D, Glennon M, Macbride-Stewart G, Moore T. Yaws in the Solomon Islands. J Trop Med Hyg. 1990;93: 52–57.

14. Chi K-H, Danavall D, Taleo F, Pillay A, Ye T, Nachamkin E, et al. Molecular Differentiation of Treponema pallidum Subspecies in Skin Ulceration Clinically Suspected as Yaws in Vanuatu Using Real-Time Multiplex PCR and Serological Methods. Am J Trop Med Hyg. 2015;92: 134–138. doi:10.4269/ajtmh.14-0459

15. Ghinai R, El-Duah P, Chi K-H, Pillay A, Solomon AW, Bailey RL, et al. A Cross-Sectional Study of ‘Yaws’ in Districts of Ghana Which Have Previously Undertaken Azithromycin Mass Drug Administration for Trachoma Control. PLoS Negl Trop Dis. 2015;9: e0003496. doi:10.1371/journal.pntd.0003496

16. Marks M, Mitjà O, Vestergaard LS, Pillay A, Knauf S, Chen C-Y, et al. Challenges and key research questions for yaws eradication. Lancet Infect Dis. 2015;15: 1220–1225. doi:10.1016/S1473-3099(15)00136-X

17. Ayove T, Houniei W, Wangnapi R, Bieb SV, Kazadi W, Luke L-N, et al. Sensitivity and specificity of a rapid point-of-care test for active yaws: a comparative study. Lancet Glob Health. 2014;2: e415–e421. doi:10.1016/S2214-109X(14)70231-1

18. Fitzpatrick C, Asiedu K, Sands A, Pena TG, Marks M, Mitja O, et al. The cost and cost-effectiveness of rapid testing strategies for yaws diagnosis and surveillance. PLoS Negl Trop Dis. 2017;11: e0005985. doi:10.1371/journal.pntd.0005985

19. Marks M, Goncalves A, Vahi V, Sokana O, Puiahi E, Zhang Z, et al. Evaluation of a Rapid Diagnostic Test for Yaws Infection in a Community Surveillance Setting. PLoS Negl Trop Dis. 2014;8: e3156. doi:10.1371/journal.pntd.0003156

20. Marks M, Yin Y-P, Chen X-S, Castro A, Causer L, Guy R, et al. Metaanalysis of the Performance of a Combined Treponemal and Nontreponemal Rapid Diagnostic Test for Syphilis and Yaws. Clin Infect Dis. 2016;63: 627–633. doi:10.1093/cid/ciw348

21. Orle KA, Gates CA, Martin DH, Body BA, Weiss JB. Simultaneous PCR detection of Haemophilus ducreyi, Treponema pallidum, and herpes simplex virus types 1 and 2 from genital ulcers. J Clin Microbiol. 1996;34: 49–54.

22. Chen C-Y, Ballard RC. The Molecular Diagnosis of Sexually Transmitted Genital Ulcer Disease. In: MacKenzie CR Henrich, editors. Diagnosis of Sexually Transmitted Diseases - Methods and Protocols. 2012. pp. 103–112.

23. Mitjà O, Lukehart SA, Pokowas G, Moses P, Kapa A, Godornes C, et al. Haemophilus ducreyi as a cause of skin ulcers in children from a yaws-endemic area of Papua New Guinea: A prospective cohort study. Lancet Glob Health. 2014;2. doi:10.1016/S2214-109X(14)70019-1

24. Marks M, Fookes M, Wagner J, Butcher R, Ghinai R, Sokana O, et al. Diagnostics for Yaws Eradication: Insights From Direct Next-Generation Sequencing of Cutaneous Strains of Treponema pallidum. Clin Infect Dis. 2018;66: 818–824. doi:10.1093/cid/cix892

25. Marks M, Chi K-H, Vahi V, Pillay A, Sokana O, Pavluck A, et al. Haemophilus ducreyi Associated with Skin Ulcers among Children, Solomon Islands. Emerg Infect Dis. 2014;20:1705–1707. doi:10.3201/eid2010.140573

26. González-Beiras C, Marks M, Chen CY, Roberts S, Mitjà O. Epidemiology of Haemophilus ducreyi Infections. Emerg Infect Dis. 2016;22: 1–8. doi:10.3201/eid2201.150425

27. Marks M, Mitjà O, Bottomley C, Kwakye C, Houinei W, Bauri M, et al. Comparative efficacy of low-dose versus standard-dose azithromycin for patients with yaws: a randomised non-inferiority trial in Ghana and Papua New Guinea. Lancet Glob Health. 2018;6: e401–e410. doi:10.1016/S2214-109X(18)30023-8

28. Palmer HM, Higgins SP, Herring AJ, Kingston MA. Use of PCR in the diagnosis of early syphilis in the United Kingdom. Sex Transm Infect. 2003;79: 479–483. doi:10.1136/sti.79.6.479

29. Mitjà O, Godornes C, Houinei W, Kapa A, Paru R, Abel H, et al. Re-emergence of yaws after single mass azithromycin treatment followed by targeted treatment: a longitudinal study. Lancet Lond Engl. 2018;391:1599–1607. doi:10.1016/S0140-6736(18)30204-6

30. Heymans R, Helm JJ van der, Vries HJC de, Fennema HSA, Coutinho RA, Bruisten SM. Clinical Value of Treponema pallidum Real-Time PCR for Diagnosis of Syphilis. J Clin Microbiol. 2010;48: 497–502. doi:10.1128/JCM.00720-09

31. Koek AG, Bruisten SM, Dierdorp M, van Dam AP, Templeton K. Specific and sensitive diagnosis of syphilis using a real-time PCR for Treponema pallidum. Clin Microbiol Infect. 2006;12:1233–1236. doi:10.1111/j.1469-0691.2006.01566.x

